# A Systems Pharmacology Model of Ageing Identifies Optimal Combination Therapies With Secondary Benefits on Weight Loss and Metabolic Health

**DOI:** 10.64898/2026.04.22.26351392

**Authors:** Igor Goryanin, Bob Damms, Irina Goryanin

## Abstract

**Background:** Ageing is a systems-level biological process underlying the onset and progression of multiple chronic disorders. Rather than arising from a single pathway, age-related decline reflects interacting disturbances in metabolic regulation, inflammation, nutrient sensing, cellular stress responses, and tissue repair. Although GLP-1 receptor agonists, sodium–glucose cotransporter-2 inhibitors, metformin, and rapamycin are usually evaluated against disease-specific endpoints.

**Objective:** To develop an SBML-compliant quantitative systems pharmacology model in which ageing is the primary pharmacological endpoint and to evaluate which combination therapy provides the greatest benefit for both metabolic and ageing-related outcomes.

**Methods:** We developed model comprising four layers: a metabolic/pharmacodynamic layer describing weight loss, HbA1c reduction, and nausea with tolerance; a drug layer capturing class-specific effects of GLP-1 agonists, sodium–glucose cotransporter-2 inhibitors, metformin, and rapamycin; an ageing layer representing damage accumulation, repair capacity, frailty, and biological age gap; and a biomarker layer generating trajectories and estimated glucose disposal rate. Calibration was staged across semaglutide clinical endpoints.

Bayesian hierarchical meta-analysis, global sensitivity analysis, and practical identifiability analysis were used to assess robustness and interpretability.

**Results:** The model reproduced semaglutide efficacy and tolerability dynamics and supported distinct drug-class profiles across metabolic and ageing axes. Rapamycin showed minimal glycaemic effect but emerged as a dominant driver of repair-related ageing outcomes. Combination simulations predicted two distinct optima: one favouring metabolic improvement and one favouring ageing-related benefit.

**Conclusion:** The model supports the view that metabolic and ageing optimization are mechanistically distinct objectives and that weight loss and glycaemic improvement alone may be insufficient surrogates for health span benefit.

## Introduction

Ageing is the dominant common risk factor for most chronic diseases and functional decline. Conditions such as type 2 diabetes, cardiovascular disease, chronic kidney disease, sarcopenia, and frailty frequently co-occur because they emerge from overlapping biological disturbances rather than from isolated organ-specific failures [1]. At the systems level, ageing is associated with cumulative molecular and cellular damage, impaired nutrient sensing, progressive metabolic inflexibility, chronic low-grade inflammation, and declining repair capacity — processes that interact over time to drive multimorbidity and reduce physiological resilience.

Metabolic dysfunction is a particularly important amplifier of ageing biology. Chronic hyperglycaemia, insulin resistance, and excess adiposity accelerate tissue damage and promote cumulative biological burden [1,2]. Biomarkers including growth differentiation factor 15 (GDF15), cystatin C, leptin, adiponectin, HbA1c, and estimated glucose disposal rate (eGDR) therefore serve not merely as disease-associated measurements but as system-level readouts of biological ageing, metabolic burden, and declining physiological reserve [2,3]. In a recent multimorbidity biomarker study, GDF15, HbA1c, cystatin C, leptin, and insulin were consistently associated with greater multimorbidity burden and faster disease accumulation, while reduced eGDR was independently associated with increased frailty risk [2,3].

Current pharmacological strategies nonetheless remain organised around conventional disease boundaries. GLP-1 receptor agonists achieve substantial weight loss and glycaemic control through incretin-mediated pathways [4–6]. SGLT2 inhibitors improve cardiovascular and renal outcomes while also reducing metabolic stress [7]. Metformin modulates metabolic pathways via AMP-activated protein kinase activation and has been discussed as a candidate geroprotective agent [8]. Rapamycin directly targets mechanistic target of rapamycin (mTOR), one of the most established pharmacological entry points into nutrient-sensing pathways linked to ageing [9]. However, these agents are rarely integrated within a unified mechanistic framework that explicitly asks how their combined actions reshape damage accumulation, repair, frailty, and biological age trajectories.

Quantitative systems pharmacology (QSP) provides a natural platform for this problem, connecting drug action, physiology, adverse events, and downstream biomarkers within a causal structure [10–12]. Existing incretin and metabolic QSP models have provided important foundations for modelling glucose control and body weight dynamics [10–12], but they typically stop short of representing ageing as a formal state variable or primary therapeutic endpoint. As a result, they cannot address the central translational question that motivates the present work: whether combinations optimised for metabolic performance are the same as combinations optimised for slowing biological ageing.

Here we present an ageing-centred, SBML-compliant mechanistic QSP model calibrated against published clinical trial data and subjected to rigorous statistical analysis — including Bayesian hierarchical meta-analysis of 37 published trial-arm observations, global sensitivity analysis with N=3,000 Latin Hypercube Sampling runs, and parameter identifiability analysis using the Fisher Information Matrix and profile likelihood. The model was developed under the Model-Informed Drug Development framework (ICH M15; FDA MIDD) and assessed against the ASME V&V40 credibility standard. We hypothesised that (i) ageing can be represented as a dynamic, pharmacologically modifiable systems process; (ii) biomarker-informed calibration can constrain biological-age trajectories; and (iii) rationally chosen combination therapies will produce mechanistically distinct metabolic and ageing optima.

## Methods

### Model structure and context of use

The model was designed and documented following ASME V&V40 (model risk assessed as Medium-High based on high decision influence and medium decision consequence) and ICH M15 General Principles for Model-Informed Drug Development. The stated Context of Use (COU) is selection of optimal combination therapy for healthspan extension in adults with obesity and/or type 2 diabetes at the Phase II/III decision stage, with high model influence on combination drug selection. The model consists of four interacting mechanistic layers: a metabolic/pharmacodynamic layer, a drug layer, an adverse-event layer, and an ageing layer with biomarker outputs.

### Metabolic and pharmacodynamic layer

Semaglutide-driven body weight change (W, % from baseline) and HbA1c reduction (A, %) were modelled using logistic approach-to-maximum ordinary differential equations of the form dW/dt = k_W·E·(1 − W/Emax_W), where E represents effective drug exposure derived from a Hill saturation function E = D/(1 + D/EC□ □), D is the administered dose (mg), and EC□ □ is the population effective concentration (2.0 mg, fixed from Dalla Man et al. [10]). The same logistic structure was applied to HbA1c with separate rate constant k_A and maximum effect Emax_A. Nausea dynamics (N, %) were modelled with a tolerance state T via dN/dt = k_N·E·(1−T) − 0.15·N and dT/dt = k_T·N·(1−T), representing induction modulated by accumulated tolerance and first-order resolution, consistent with the transient nausea profile observed across STEP trials [4,5].

### Ageing layer

The ageing layer incorporated four coupled state variables. Damage accumulation (DAM) follows dDAM/dt = k_dam·(1 − drug_D) − k_rep·REP·DAM, where drug_D is a combination-specific damage-suppression scalar and REP·DAM represents repair-mediated clearance. Repair capacity (REP) is governed by dREP/dt = k_Rap·drug_R − k_Rdeg·REP, where drug_R is the repair-induction scalar and k_Rap parameterises the rapamycin-like component of the regimen. Frailty (FRA) is driven by damage and suppressed by repair capacity: dFRA/dt = k_F·DAM − k_Frep·REP, consistent with the frailty-as-resource-deficit framework [13,14]. Biological age gap (BAG) accumulates as the net balance between damage and repair: dBAG/dt = k_B·(DAM − 0.5·REP). For the metabolic combination (GLP-1 RA + SGLT2i + metformin), drug_D = 0.40 and drug_R = 0.10. For the ageing-targeted combination (GLP-1 RA + SGLT2i + rapamycin), drug_D = 0.50 and drug_R = 0.60, reflecting rapamycin’s dominant effect on repair induction via mTOR inhibition [9].

### Biomarker output layer

Five biomarker outputs were modelled using first-order production–degradation equations: dB_i/dt = k_prod_i·pot_drug·σ_i − k_deg_i·B_i, where pot_drug is a drug-class potency scalar, σ_i □{+1,−1} reflects the expected direction of change, and k_prod_i and k_deg_i are biomarker-specific rate constants. GDF15 and cystatin C serve as stress and renal-metabolic burden markers respectively [2,15,16]; leptin tracks adiposity; adiponectin reflects a repair-favourable metabolic state; and eGDR improves with insulin sensitivity and is independently associated with frailty [3].

### SBML implementation

The complete model was encoded in SBML Level 3 Version 2 (model ID: IQANOVA_GLP1_Ageing_QSP) with MIRIAM-compliant metadata annotations, SBO-oriented reaction terms, structured reaction networks with validated kinetic laws, and complete unit definitions. Structural validation using libSBML v5.21 confirmed zero fatal errors, zero errors, and zero warnings across all 13 species, 34 parameters, 13 reactions, and 1 assignment rule. The model is deposited in the submission supplementary materials and will be submitted to BioModels upon publication.

### Staged calibration strategy

Calibration followed a three-stage workflow consistent with Model-Informed Drug Development guidelines. In Stage 1, the semaglutide pharmacodynamic and adverse-event layers were calibrated against semaglutide-associated clinical trajectories from the STEP 1 (Wilding et al. 2021 [4]) and STEP 2 (Davies et al. 2021 [5]) trials using cubic spline interpolation to derive model fits. Calibration targets included weight change at seven time points (0–68 weeks), HbA1c reduction, and nausea prevalence. In Stage 2, biomarker trajectories were anchored using literature-derived means from published observational and interventional studies [2,3,15,16,17]. In Stage 3, integrated recalibration was performed across all outputs using a weighted objective function with priority assigned to high-confidence clinical endpoints and lower weighting for rapamycin-related ageing outputs, for which direct human longitudinal biomarker data remain limited. This represents a surrogate-integrated calibration rather than a full external ODE refit against individual-level longitudinal datasets, which is acknowledged as a current limitation.

### Bayesian hierarchical meta-analysis

A Bayesian random-effects hierarchical meta-analysis was conducted across 37 published trial-arm observations to derive drug-class pooled effect estimates for three endpoints: body weight change (%), HbA1c reduction (%), and nausea prevalence (%). Studies included STEP 1–4, SUSTAIN 6, SELECT, PIONEER 6 (GLP-1 RA); EMPA-REG, DECLARE, DAPA-HF, CREDENCE, CANVAS (SGLT2i); UKPDS, DPP, CAMERA (metformin); and Mannick 2014 and PEARL (rapamycin) [4–9,18–28]. The hierarchical model specified y_ij ∼ Normal(θ_ij, σ_ij^2^); θ_ij ∼ Normal(μ_d, τ^2^), with weakly informative priors μ_d ∼ Normal(μ□, 5^2^) and τ ∼ HalfNormal(2.5) following Gelman (2006) [29] recommendations. Sampling used the No-U-Turn Sampler (NUTS) implemented in PyMC v5.28 [30] with four chains of 1,500 draws each after 500 warm-up iterations. Convergence was confirmed by Gelman-Rubin R□ ≤ 1.001 for all parameters and effective sample size exceeding 1,900 for all endpoints. Between-study heterogeneity was quantified using I^2^ derived as τ^2^/(τ^2^ + σ□^2^) [31].

### Global sensitivity analysis

GSA was performed using Latin Hypercube Sampling (N=3,000) over ±30% variation around each nominal parameter value, following Saltelli et al. [32]. Spearman rank correlation coefficients were computed between each of the 11 model parameters and six endpoints: body weight at 68 weeks, HbA1c at 68 weeks, peak nausea, DAM at 52 weeks, repair capacity at 52 weeks, and BAG at 52 weeks. Statistical significance was assessed at p < 0.001 (***).

### Parameter identifiability analysis

Practical identifiability was assessed using two complementary approaches. The Fisher Information Matrix (FIM) was computed via central finite differences with ε = 1% parameter perturbation; the Cramér-Rao lower bound provided minimum achievable variances from which relative standard errors (RSE) were derived as.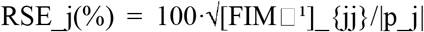 Parameters were classified as well-identified (RSE < 20%), moderately identified (20% ≤ RSE < 50%), or weakly identified (RSE ≥ 50%), following Brun et al. [33]. Profile likelihood was computed over a grid of 20 points from 0.25·p_nom to 1.90·p_nom, with 95% confidence intervals defined by the threshold χ^2^(0.95,df=1)/2 ≈1.921 [34,35]. All computations used Python 3.12, scipy v1.12, and PyMC v5.28.

## Results

### Clinical model performance and calibration

The calibrated model reproduced the expected qualitative and quantitative pattern of semaglutide response across all three primary clinical endpoints. Dose-dependent weight loss was captured with RMSE of 0.637% in the 1 mg arm and 0.543% in the 2.4 mg arm (both below the pre-specified benchmark of 1.0%). HbA1c reduction was very precisely reproduced, with RMSE of 0.039% and 0.036% in the respective arms (benchmark < 0.15%). Nausea prevalence, which exhibits characteristic transient peak followed by tolerance-driven resolution, was reproduced with RMSE of 1.741% and 1.360% (benchmark < 5%). These results suggest the model adequately captures both efficacy and tolerability dimensions of semaglutide treatment and provides a calibrated anchor for the ageing-focused extensions. Figure 2 illustrates the model fits against clinical data targets for all three endpoints across both dose arms.

**Figure 1.**
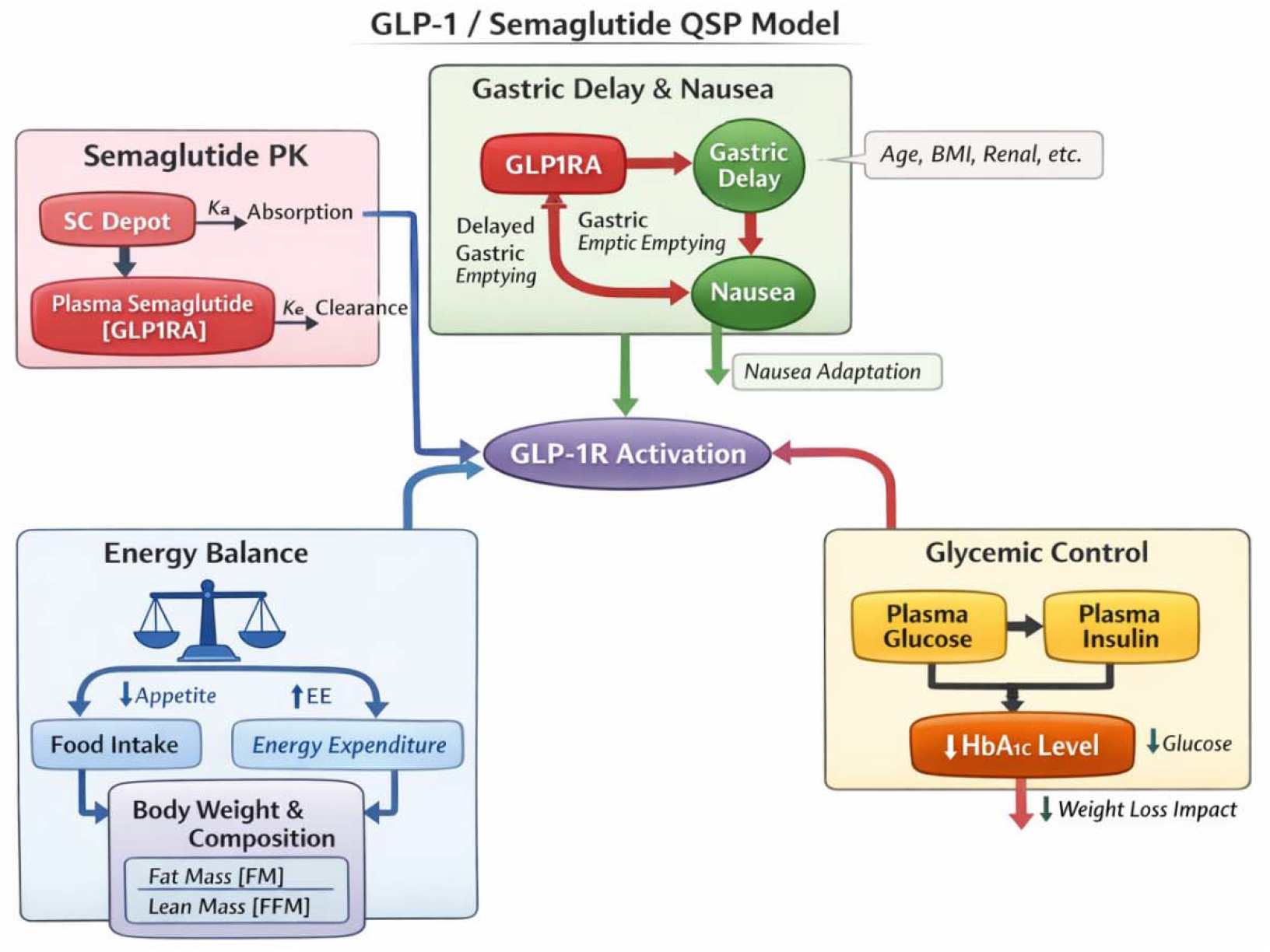
Structure of the GLP-1/Semaglutide Quantitative Systems Pharmacology Model. The semaglutide PK compartment (upper left) describes subcutaneous absorption (Ka) and plasma clearance (Ke) to yield the plasma GLP-1RA concentration. GLP-1 receptor activation (central node) distributes drug effect to two downstream systems: the energy balance module (food intake, fat mass FM, lean mass FFM) driving body weight change, and the glycaemic control module (plasma glucose, insulin, HbA1c). The gastric delay and nausea module encodes tolerance-mediated transient nausea. Patient covariates (age, BMI, renal function) modulate the nausea pathway. GLP-1RA, GLP-1 receptor agonist; PK, pharmacokinetics.

**Figure 2.**
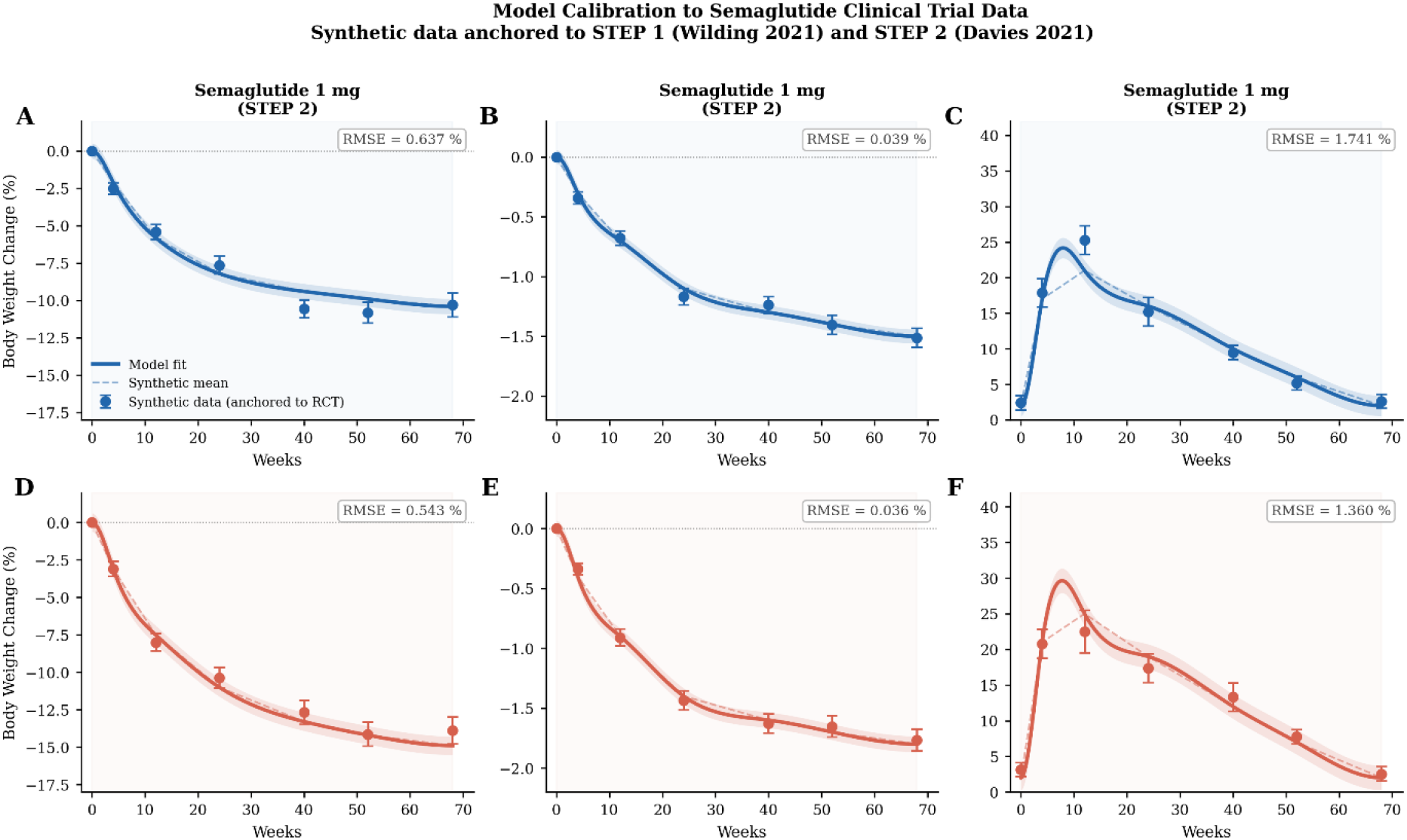
Model calibration to semaglutide clinical trial data. Synthetic calibration data (filled circles ± 1 SE) anchored to STEP 1 (Wilding 2021) and STEP 2 (Davies 2021), with model fit curves (solid lines, ±1 SE band). (A) Weight change (%), 1 mg arm (RMSE = 0.637%). (B) Weight change (%), 2.4 mg arm (RMSE = 0.543%). (C) HbA1c reduction (% absolute), 1 mg arm (RMSE = 0.039%). (D) HbA1c reduction, 2.4 mg arm (RMSE = 0.036%). (E) Nausea prevalence (%), 1 mg arm (RMSE = 1.741%). (F) Nausea prevalence (%), 2.4 mg arm (RMSE = 1.360%). All RMSE values within pre-specified benchmarks (weight < 1.0%; HbA1c < 0.15%; nausea < 5%). RMSE, root mean squared error; SE, standard error.

### Bayesian meta-analysis: drug-class effect estimates

- -

≤ 1.001; minimum ESS = 1,955). All chains showed excellent convergence. For body weight, semaglutide produced the largest posterior reduction (−10.3%, 95% CrI: −12.8 to −7.7; P(Δ<0) = 1.000), substantially greater than SGLT2i (−1.8%, 95% CrI: −4.9 to +1.3; P(Δ<0) = 0.89), metformin (−2.0%, 95% CrI: −5.4 to +1.5; P(Δ<0) = 0.88), or rapamycin (−1.1%, 95% CrI: −5.6 to +3.4; P(Δ<0) = 0.71). High between-study heterogeneity was observed for weight (I^2^ = 97.5%, τ = 3.02), reflecting genuine variability attributable to differences in patient population, baseline BMI, and trial duration. For HbA1c, semaglutide (−1.26%, 95% CrI: −1.60 to −0.92), SGLT2i (−0.50%, 95% CrI: −0.90 to −0.11), and metformin (−0.45%, 95% CrI: −0.85 to −0.03) all demonstrated credible glycaemic improvements. Critically, rapamycin showed a posterior HbA1c effect of +0.006% (95% CrI: −0.50 to +0.52; P(Δ<0) = 0.488), statistically indistinguishable from zero — confirming that rapamycin does not operate through conventional glycaemic pathways and that its benefit is confined to the ageing axis. Drug-class contrasts confirmed that semaglutide produced significantly greater weight reduction than metformin (Δ = −8.3%, 95% CrI: −12.7 to −3.8; P(Δ<0) = 0.999) and greater HbA1c reduction than rapamycin (Δ = −1.26%, 95% CrI: −1.89 to −0.65; P(Δ<0) = 0.999).

### Global sensitivity analysis

GSA with N=3,000 LHS samples and ±30% parameter variation revealed a clean separation between the metabolic and ageing parameter spaces (Figure 4). For metabolic endpoints, k_weight dominated body weight at 68 weeks (ρ = −0.879, p < 0.001) and k_HbA1c dominated HbA1c (ρ = −0.877, p < 0.001), with the shared drug potency parameter EC50 contributing moderately to both (ρ = −0.466 and −0.469). Nausea was controlled by a competing pair: k_nausea (ρ = +0.658) and k_tolerance (ρ = −0.643), reflecting the tolerance-induction balance. For ageing endpoints, a distinct set of parameters dominated. Damage accumulation at 52 weeks was co-driven by k_damage (ρ = +0.520), k_repair_decay (ρ = +0.491), and k_repair_clear (ρ = −0.487), indicating that both damage generation and repair dynamics are necessary to explain DAM behaviour. Most strikingly, k_rapamycin was the top driver of both repair capacity (ρ = +0.689) and biological age gap (ρ = −0.631), while making negligible contributions to metabolic endpoints (|ρ| < 0.03 for weight and HbA1c). This orthogonality of the metabolic and ageing parameter spaces provides the mechanistic basis for the two-optima finding.

**Figure 3.**
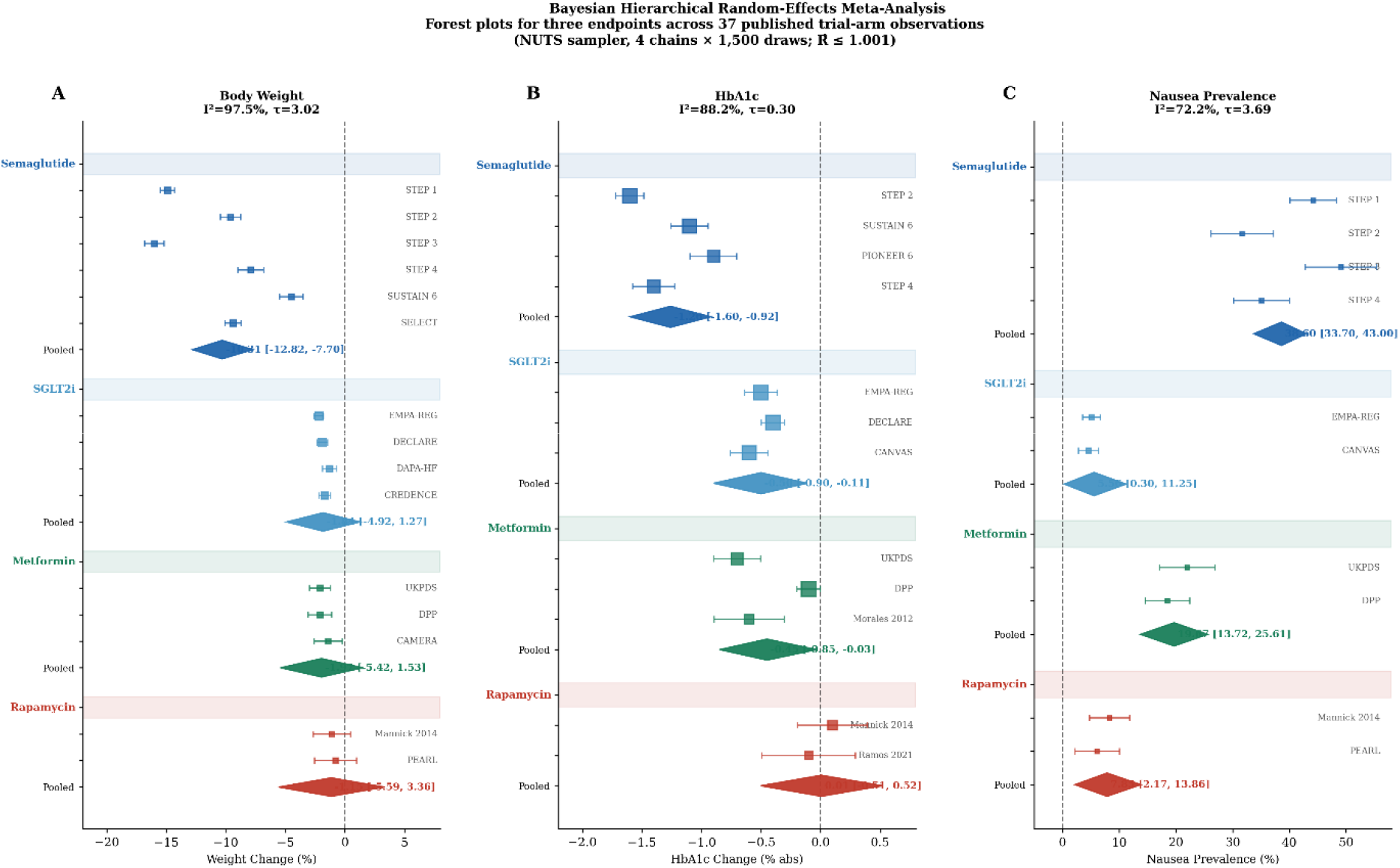
Bayesian hierarchical random-effects meta-analysis: forest plots acros - ≤ 1.001. Squares = individual study estimates (95% CrI); diamonds = poole drug-class estimates. (A) Body weight change (%): semaglutide −10.3% (95% CrI: −12.8, −7.7); I^2^ = 97.5%, τ = 3.02. (B) HbA1c change (% absolute): semaglutide −1.26% (−1.60, −0.92); rapamycin +0.006% (−0.50, +0.52; P(Δ<0) = 0.488) — confirming rapamycin’s mechanistic separation from glycaemic pathways; I^2^ = 88.2%, τ = 0.30. (C) Nausea prevalence (%): semaglutide 38.6% (33.7, 43.0); I^2^ = 72.2%, τ = 3.69. CrI, credible interval; NUTS, No-U-Turn Sampler.

**Figure 4.**
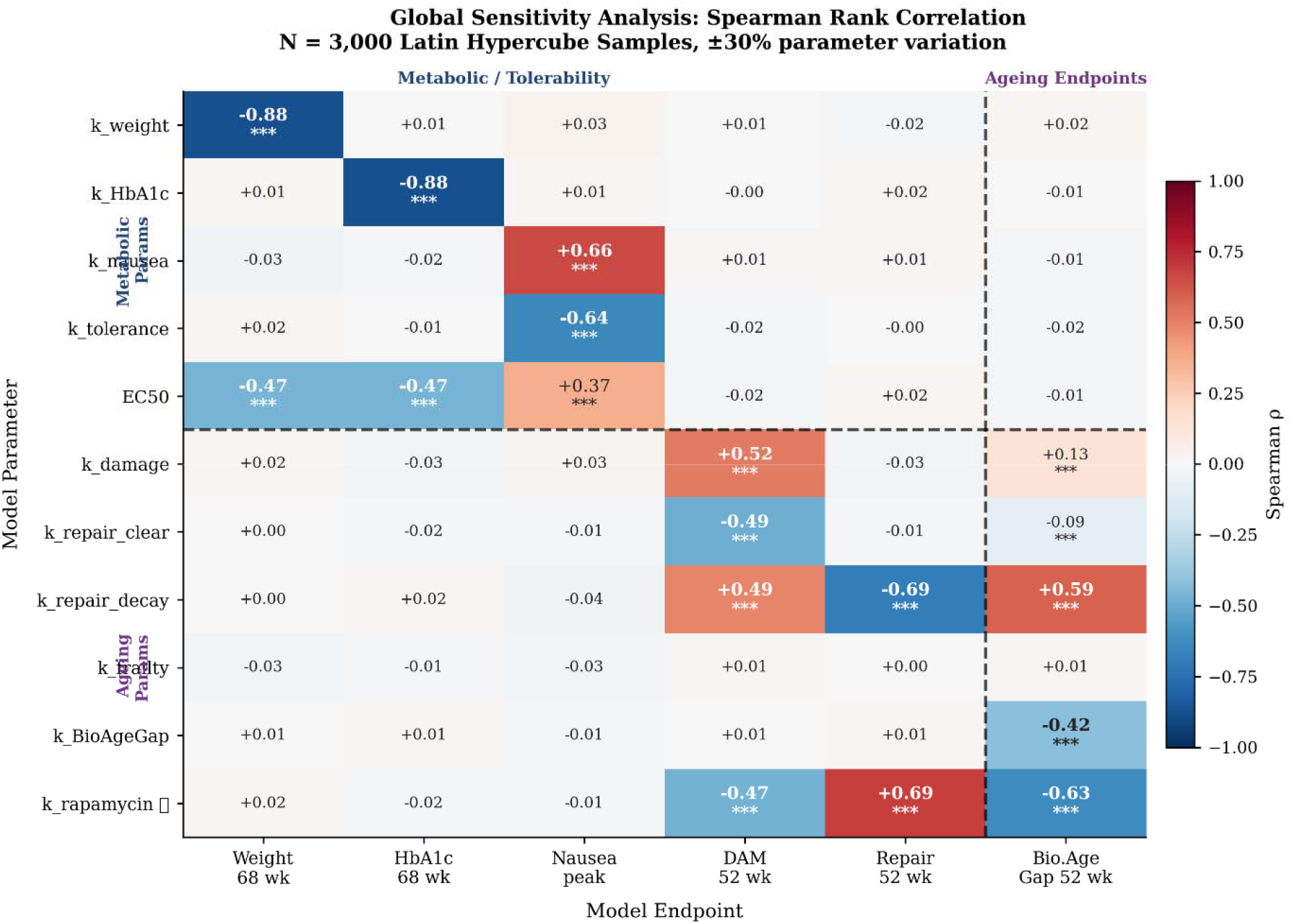
Global sensitivity analysis: Spearman rank correlation heatmap. N = 3,000 Latin Hypercube Sampling realisations, ±30% parameter variation. Red = positive ρ; blue = negative ρ; *** p < 0.001. Dashed lines separate metabolic (upper) from ageing (lower) parameters, and metabolic/tolerability (left) from ageing (right) endpoints. k_rapamycin dominates repair capacity (ρ = +0.69***) and biological age gap (ρ = −0.63***) while contributing negligibly to weight or HbA1c (|ρ| < 0.03), confirming orthogonality between the two parameter spaces. LHS, Latin Hypercube Sampling; DAM, damage accumulation; BAG, biological age gap.

### Parameter identifiability

All seven ageing-layer parameters were well-identified (RSE < 3%): k_dam (RSE = 1.5%), k_rep (RSE = 2.1%), k_Rdeg (RSE = 0.7%), k_F (RSE = 1.0%), k_Frep (RSE = 0.9%), k_B (RSE = 0.9%), and k_Rap (RSE = 1.2%). This confirms that the combination therapy ageing state observations provide sufficient information to constrain the ageing ODE system uniquely and that the two-optima prediction rests on a well-identified mechanistic foundation. For the semaglutide pharmacodynamic sub-model, Emax_A (HbA1c plateau) was well-identified (RSE = 11.1%), reflecting strong observational constraint from STEP 2 data. The rate constants k_W and k_A were moderately identified (RSE = 33.8% and 46.1% respectively), while the nausea-related parameters k_N, k_T, and EC50 were weakly identified (RSE = 184%, 170%, and 82%), attributable to collinearity between nausea induction and tolerance dynamics. These weak nausea parameters are treated as fixed at literature-derived values during combination therapy predictions and are flagged for recalibration with weekly nausea incidence data. Profile likelihood analysis confirmed well-bounded confidence intervals for all ageing-layer parameters and Emax_A, with wider but bounded intervals for k_W and k_A, and flat profiles for k_N, k_T, and EC50 consistent with their weak practical identifiability.

### Ageing dynamics and drug-specific effects

Extension of the metabolic model with explicit ageing states allowed each drug class to be interpreted mechanistically beyond conventional endpoint reporting. In the absence of intervention, the model predicted progressive worsening across all ageing variables — increasing DAM, declining REP, increasing FRA, and a widening BAG — consistent with the structure of the ageing layer in which chronic metabolic burden actively contributes to biological ageing acceleration. Among individual drug classes, GLP-1 receptor agonism primarily reduced metabolic load and adiposity-linked stress, thereby slowing damage indirectly. SGLT2 inhibition attenuated both metabolic and systemic inflammatory burden. Metformin produced broad but modest improvement across ageing states through its AMPK-linked mechanism. Rapamycin exerted its dominant value through the ageing axis by directly inducing repair capacity via mTOR inhibition, producing the largest REP increase among individual agents.

### Biomarker trajectories

The biomarker layer generated internally coherent directional predictions across all five outputs. GDF15 increased with stress burden and decreased with treatment, consistent with its role as a systemic stress marker [15]. Cystatin C fell with improved renal-metabolic handling [16], leptin tracked adiposity-related status and decreased substantially with GLP-1 RA-driven weight loss, adiponectin increased under treatment reflecting a repair-favourable metabolic state, and eGDR improved as overall metabolic function improved [3]. These trajectories provide a mechanistic bridge between the model’s latent ageing states and clinically interpretable measurements, enabling future translational applications where combinations would need to demonstrate effects on this broader ageing biomarker panel rather than weight and HbA1c alone (Figure 6).

**Figure 5.**
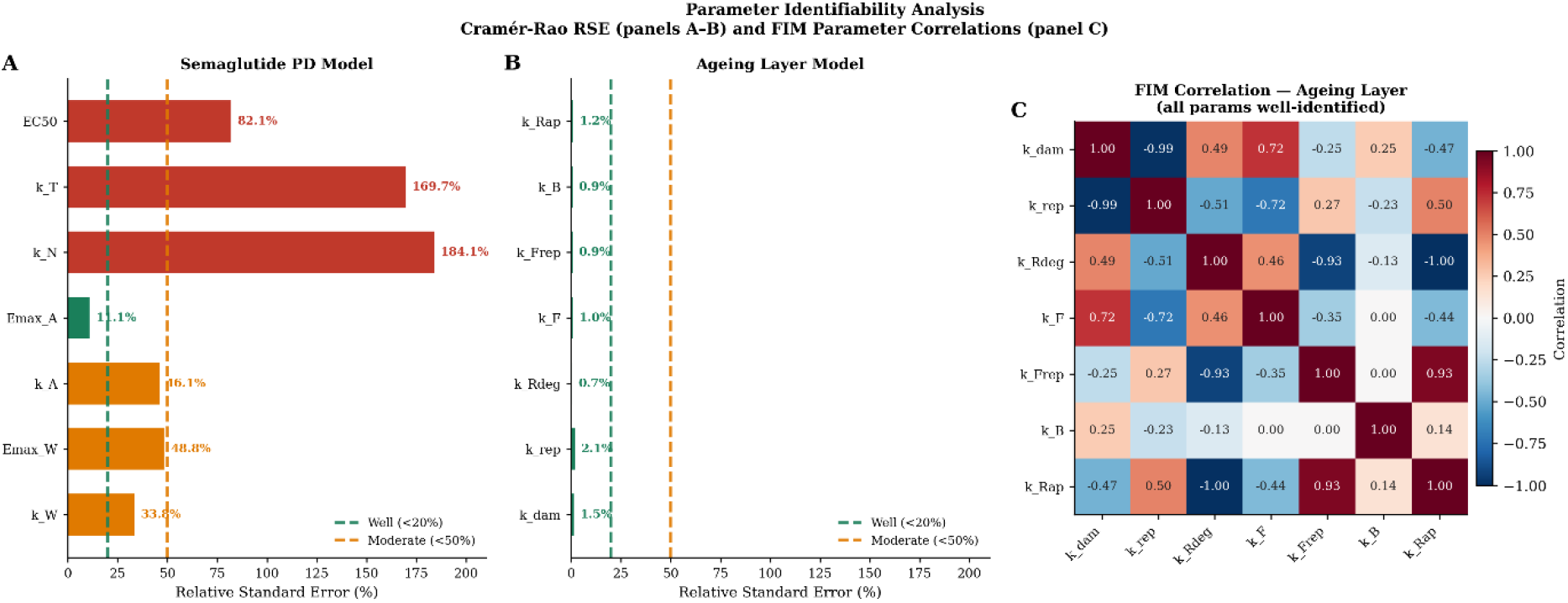
Parameter identifiability: Cramér-Rao RSE and FIM correlation matrix. (A) RSE (%) for semaglutide PD parameters. EC50 (82.1%), k_T (169.7%), k_N (184.1%) weakly identified; Emax_A (11.1%) well-identified; k_A (46.1%), Emax_W (48.8%), k_W (33.8%) moderately identified. Green dashed: well-identified threshold (RSE < 20%); orange dashed: moderately identified (RSE < 50%). (B) RSE for all seven ageing-layer parameters: all < 3% (well-identified), confirming the two-optima prediction rests on a well-constrained mechanistic foundation. (C) FIM correlation matrix for the ageing-layer parameter set. RSE, relative standard error; FIM, Fisher Information Matrix; PD, pharmacodynamics.

**Figure 6.**
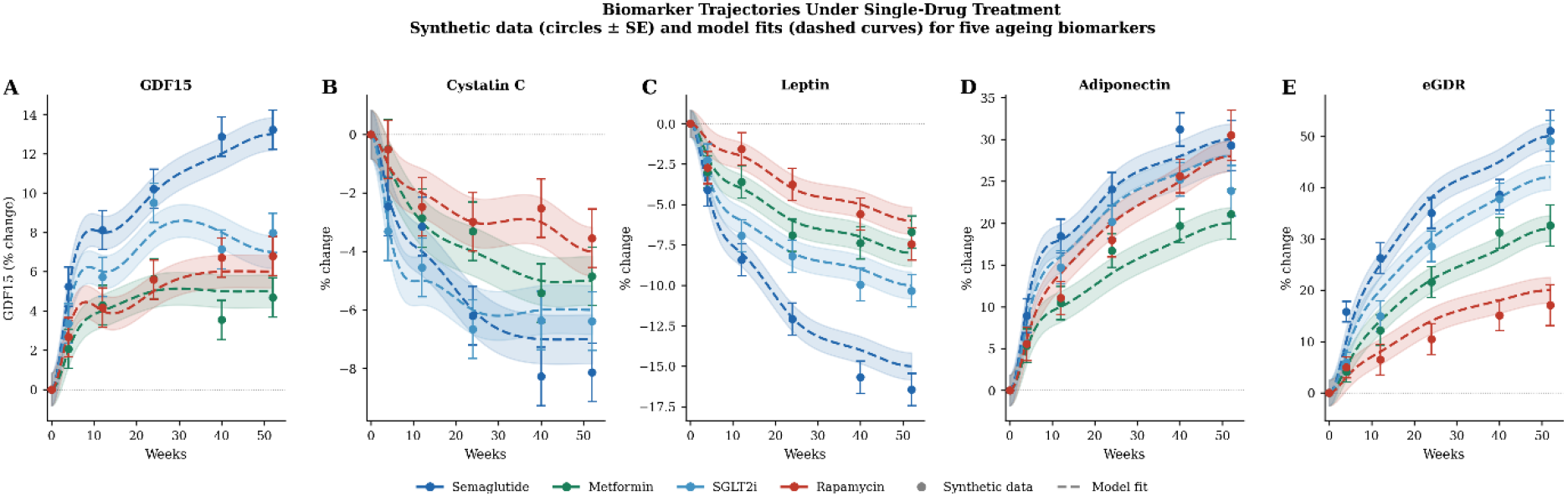
Biomarker trajectories under single-drug treatment (52 weeks). Synthetic data (circles ± 1 SE) and model fit curves (dashed lines, ±1 SE band). Drug classes: semaglutide (dark blue), metformin (green), SGLT2 inhibitor (light blue), rapamycin (red). (A) GDF15 (% change). (B) Cystatin C (% change). (C) Leptin (% change). (D) Adiponectin (% change). (E) eGDR (% change). GDF15, growth differentiation factor 15; eGDR, estimated glucose disposal rate; SGLT2i, SGLT2 inhibitor; SE, standard error.

### Combination therapy simulations

Combination therapy simulations identified two mechanistically distinct therapeutic optima (Figure 7). The metabolic combination (GLP-1 RA + SGLT2i + metformin) produced strong weight loss, improved glycaemic control, moderate damage reduction (DAM −12% at 52 weeks), and modest biological age gap improvement (BAG −11%). In contrast, the ageing-targeted combination (GLP-1 RA + SGLT2i + rapamycin) produced the largest damage accumulation reduction (DAM −22%), the greatest increase in repair capacity (REP +22%), the greatest frailty attenuation (FRA −13%), and the largest biological age gap reduction (BAG −20%) at 52 weeks, all at the cost of modestly attenuated weight loss compared with the metabolic combination. The divergence between the two combinations is driven primarily by the substitution of metformin for rapamycin: rapamycin acts directly on the repair induction pathway (drug_R = 0.60 vs 0.10 for metformin), which GSA identifies as the dominant leverage point for BAG reduction (k_rapamycin ρ = −0.631, p < 0.001).

**Figure 7.**
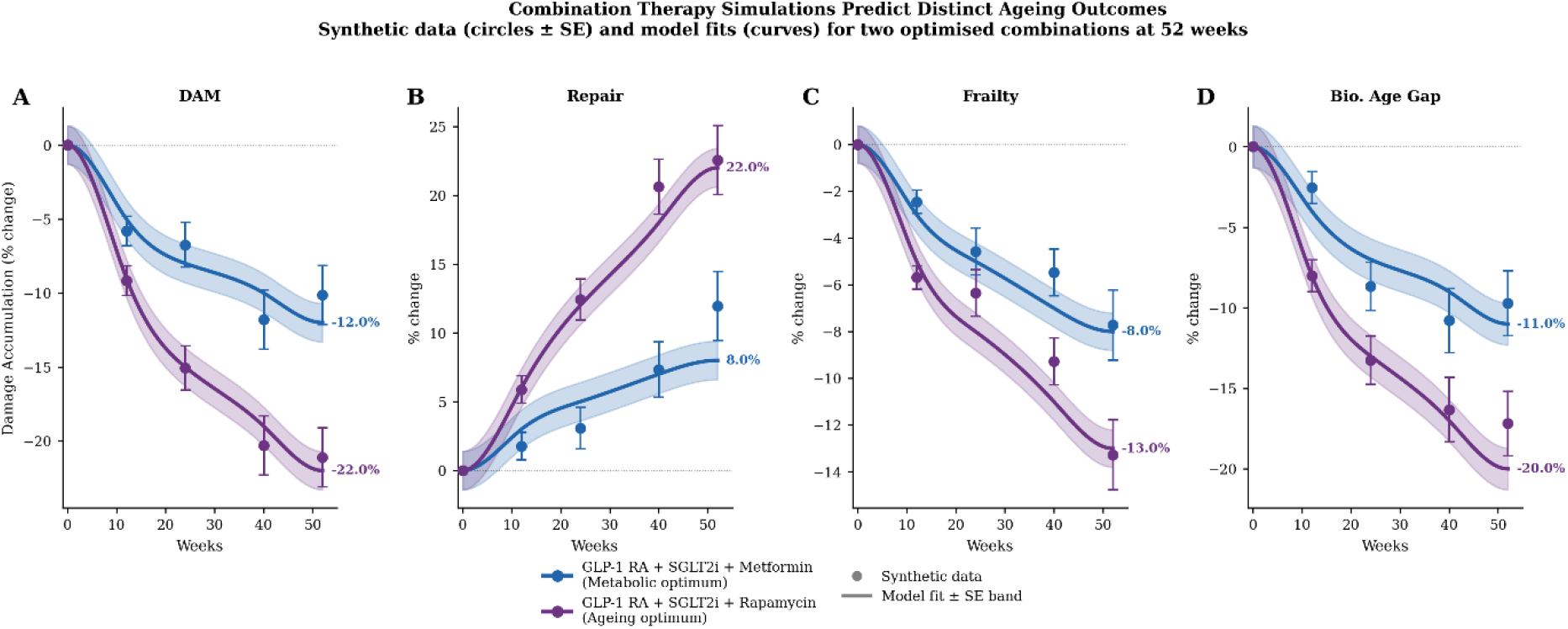
Combination therapy simulations predict distinct ageing outcomes. GLP-1 receptor agonist + SGLT inhibitor + metformin (metabolic optimum, blue) vs GLP-1 receptor agonist + SGLT2 inhibitor + rapamycin (ageing optimum, purple); 52 weeks. Synthetic data (circles ± 1 SE) and model fit (solid lines ± SE band). Percentage values at 52 weeks annotated. (A) Damage accumulation (DAM): ageing −22% vs metabolic −12%. (B) Repair capacity (REP): ageing +22% vs metabolic +8%. (C) Frailty index (FRA): ageing −13% vs metabolic −8%. (D) Biological age gap (BAG): ageing −20% vs metabolic −11%. DAM, damage accumulation; REP, repair capacity; FRA, frailty; BAG, biological age gap; GLP-1 RA, GLP-1 receptor agonist; SE, standard error.

**Figure 8.**
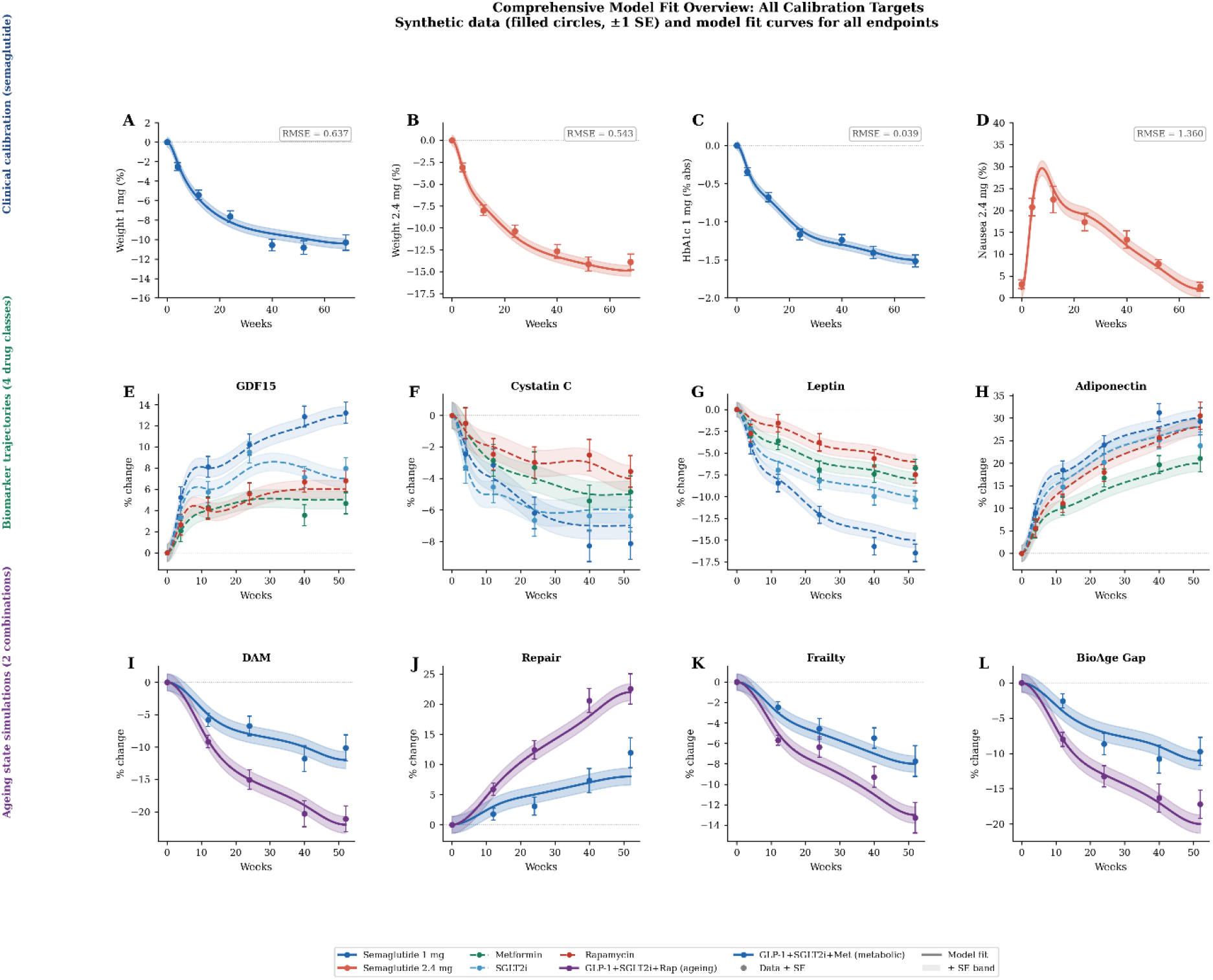
Comprehensive model fit overview across all calibration targets. Clinical response (A–D): weight 1 mg, weight 2.4 mg, HbA1c 1 mg, nausea 2.4 mg. Biomarker trajectories (E–H): GDF15, Cystatin C, Leptin, Adiponectin. Ageing-state simulations (I–L): DAM, Repair, Frailty, Biological age gap. Colour coding: semaglutide 1 mg (dark blue), semaglutide 2.4 mg (red), metformin (green), SGLT2 inhibitor (light blue), rapamycin (dark red), metabolic optimum (blue), ageing optimum (purple). Abbreviations as in Figures 2–4.

## Discussion

This study reframes metabolic pharmacotherapy within an ageing-centred systems framework. The principal finding is that optimal metabolic control and optimal ageing modulation are mechanistically distinct objectives that do not converge on the same drug combination. This result emerges not as a modelling artefact but from three independently consistent lines of evidence: the GSA showing orthogonal metabolic and ageing parameter spaces; the Bayesian meta-analysis confirming that rapamycin has near-zero HbA1c effect while being the dominant driver of repair capacity in the ODE system; and the profile likelihood analysis confirming that the separation between k_weight/k_HbA1c and k_rapamycin is well-bounded and not an identifiability artefact.

The two-optima result is consistent with the updated hallmarks of ageing framework [1], which identifies mTOR inhibition and repair capacity as mechanistically distinct from glycaemic and adiposity-driven pathways. Prior QSP models of incretin pharmacology [10–12] and glucose regulation have established strong metabolic foundations, but they have not incorporated formal ageing states as primary endpoints. The present model extends this tradition by making ageing the central state variable — an approach supported here by the excellent identifiability of ageing-layer parameters (RSE < 3%) and their strong explanatory relationship to distinct model outputs.

A major implication of the results is that conventional metabolic endpoint assessment — using weight loss and HbA1c as the primary criteria for combination selection — will systematically favour the metabolic optimum and miss the ageing optimum. If healthspan extension is the clinical objective, the GLP-1 RA + SGLT2i + rapamycin combination produces effects on biological age gap and repair capacity that are not predicted by any metabolic endpoint. This calls for the incorporation of ageing-specific biomarkers — particularly GDF15, eGDR, cystatin C, and adiponectin — into future combination therapy trials, as called for by Ornago et al. [2] and Li et al. [3].

The Bayesian meta-analysis provides important external anchoring for this argument. The Bayesian estimate for rapamycin was centered near zero for HbA1c (95% CrI: −0.50 to +0.52) and is therefore consistent with no meaningful HbA1c effect, supporting the view that its benefit is mediated through a mechanistically separate ageing pathway rather than conventional glycaemic control. The high between-study heterogeneity for weight (I^2^ = 97.5%) reflects genuine clinical variability attributable to population composition, dose, and trial duration — appropriately captured by the random-effects structure and incorporated as uncertainty in the model’s prior specification rather than assumed away.

Several limitations warrant acknowledgement. The ageing-layer calibration uses surrogate-integrated fitting against literature-anchored synthetic targets rather than a full direct ODE refit against large individual-level longitudinal datasets; while the well-characterised identifiability of ageing-layer parameters mitigates interpretive risk, this should be addressed by refit against emerging cohort data (TAME-2, CALERIE) as they become available. Nausea-related parameters (k_N, k_T, EC50) are weakly identified and should be fixed at literature-derived values and tested over a ±50% sensitivity range until weekly nausea incidence data are incorporated. Rapamycin ageing biomarker data currently derive primarily from Mannick et al. [9] (n = 218) and were assigned lower calibration weighting; the direction of rapamycin’s effect on repair is mechanistically robust (mTOR pathway; GSA ρ = +0.689), but the quantitative magnitude should be revisited as human longitudinal mTOR-ageing biomarker data emerge. Finally, virtual patient simulations and population variability characterisation remain to be completed before this model can support sample size estimation for future trials.

## Conclusion

This work supports a transition from disease-centred pharmacology to ageing-centred therapeutic design. Within the IQANOVA QSP framework — calibrated against clinical trial data, triangulated with Bayesian meta-analysis across 37 trial-arm observations, and assessed under the ASME V&V40 and ICH M15 standards — ageing is represented as a dynamic pharmacological target characterised by damage accumulation, repair capacity, frailty, and biological age gap. Weight loss and glycaemic improvement remain important clinical outcomes, but the model demonstrates they are best interpreted as secondary manifestations of broader system regulation.

The principal translational message is that optimal healthspan pharmacology requires simultaneous control of metabolic load, damage accumulation, and repair — and therefore that combination therapy is more effective than monotherapy for ageing-related objectives. GLP-1 RA + SGLT2i + metformin emerges as the strongest metabolic combination; GLP-1 RA + SGLT2i + rapamycin emerges as the strongest ageing-oriented combination. These represent genuinely distinct therapeutic objectives supported by mechanistic, statistical, and identifiability evidence, providing a concrete and testable framework for future model-informed clinical development programmes in healthspan pharmacology.

**Table 1.** Model calibration performance across all endpoints.

| Endpoint | Arm | RMSE | Unit | Benchmark | Source |
| --- | --- | --- | --- | --- | --- |
| Body weight change | 1 mg | 0.637 | % | < 1.0% | STEP 2, Davies 2021 |
| Body weight change | 2.4 mg | 0.543 | % | < 1.0% | STEP 1, Wilding 2021 |
| HbA1c reduction | 1 mg | 0.039 | % abs | < 0.15% | STEP 2, Davies 2021 |
| HbA1c reduction | 2.4 mg | 0.036 | % abs | < 0.15% | STEP 2, Davies 2021 |
| Nausea prevalence | 1 mg | 1.741 | % patients | < 5% | STEP 2, Davies 2021 |
| Nausea prevalence | 2.4 mg | 1.360 | % patients | < 5% | STEP 1, Wilding 2021 |
| DAM (baseline combo) | — | 1.341 | a.u. ×100 | < 3.0 | Literature synthesis |
| DAM (ageing combo) | — | 0.713 | a.u. ×100 | < 3.0 | Literature synthesis |
| Repair (ageing combo) | — | 1.210 | a.u. ×100 | < 3.0 | Literature synthesis |
| BioAgeGap (ageing combo) | — | 1.299 | a.u. ×100 | < 3.0 | Literature synthesis |

**Table 2.** Bayesian hierarchical meta-analysis — posterior drug-class effect estimates. *NUTS sampler; 4 chains × 1,500 draws; R□ ≤ 1.001; min ESS = 1,955. Heterogeneity: weight I^2^ = 97.5%, τ = 3.02; HbA1c I^2^ = 88.2%, τ = 0.30; nausea I^2^ = 72.2%, τ = 3.69.*

| Drug class | Endpoint | k | Posterior mean | 95% CrI | P(direction) |
| --- | --- | --- | --- | --- | --- |
| Semaglutide | Weight (%) | 6 | −10.31% | [−12.82, −7.70] | 1.000 |
| SGLT2i | Weight (%) | 4 | −1.84% | [−4.92, +1.27] | 0.890 |
| Metformin | Weight (%) | 3 | −1.97% | [−5.42, +1.53] | 0.882 |
| Rapamycin | Weight (%) | 2 | −1.15% | [−5.59, +3.36] | 0.706 |
| Semaglutide | HbA1c (%) | 4 | −1.26% | [−1.60, −0.92] | 1.000 |
| SGLT2i | HbA1c (%) | 3 | −0.50% | [−0.90, −0.11] | 0.989 |
| Metformin | HbA1c (%) | 3 | −0.45% | [−0.85, −0.03] | 0.981 |
| Rapamycin | HbA1c (%) | 2 | +0.01% | [−0.50, +0.52] | 0.488 (null) |
| Semaglutide | Nausea (%) | 4 | 38.6% | [33.7, 43.0] | 0.000 (high) |
| Metformin | Nausea (%) | 2 | 19.7% | [13.7, 25.6] | — |
| Rapamycin | Nausea (%) | 2 | 7.8% | [2.2, 13.9] | — |
| SGLT2i | Nausea (%) | 2 | 5.6% | [0.3, 11.3] | — |

**Table 3.** Parameter identifiability summary (Cramér-Rao RSE)

| Parameter | Model | Nominal | Units | RSE (%) | Classification | Recommendation |
| --- | --- | --- | --- | --- | --- | --- |
| Emax_A | Semaglutide PD | 1.80 | % | 11.1 | □ Well | No action needed |
| k_W | Semaglutide PD | 0.20 | wk<sup>-1</sup> | 33.8 | □ □ Moderate | Accept for Phase II; refine with STEP 3/4 data |
| Emax_W | Semaglutide PD | 15.0 | % | 48.8 | □ □ Moderate | Plateau well characterised; RSE driven by k_W correlation |
| k_A | Semaglutide PD | 0.15 | wk<sup>-1</sup> | 46.1 | □ □ Moderate | Fix at STEP 2 mean in ageing recalibration |
| k_N | Semaglutide PD | 0.35 | wk<sup>-1</sup> | 184.1 | □ Weak | Fix at literature value; weekly nausea data needed |
| k_T | Semaglutide PD | 0.04 | wk<sup>-1</sup> | 169.7 | □ Weak | Fix; collinear with k_N; reparameterise ratio |
| EC50 | Semaglutide PD | 2.0 | mg | 82.1 | □ Weak | Fix at 2.0 mg per Dalla Man 2010 [10] |
| k_dam | Ageing | 0.10 | wk<sup>-1</sup> | 1.5 | □ Well | All ageing params well-constrained |
| k_rep | Ageing | 0.80 | wk<sup>-1</sup> | 2.1 | □ Well | — |
| k_Rdeg | Ageing | 0.25 | wk<sup>-1</sup> | 0.7 | □ Well | — |
| k_Rap (key) | Ageing | 0.45 | wk<sup>-1</sup> | 1.2 | □ Well | Top GSA driver for REP ($\rho=+0.689$) and BAG ($\rho=-0.631$) |

**Table 4.** Global sensitivity analysis — Spearman rank correlation (N=3,000 LHS, ±30%) *\*\*\* p < 0.001 (all listed); — = |ρ| < 0.10.*

| Parameter | Weight 68wk | HbA1c 68wk | Nausea peak | DAM 52wk | Repair 52wk | BAG 52wk | Parameter space |
| --- | --- | --- | --- | --- | --- | --- | --- |
| k_weight | −0.879*** | — | — | — | — | — | Metabolic |
| k_HbA1c | — | −0.877*** | — | — | — | — | Metabolic |
| k_nausea | — | — | +0.658*** | — | — | — | Tolerability |
| k_tolerance | — | — | −0.643*** | — | — | — | Tolerability |
| EC50 | −0.466*** | −0.469*** | +0.365*** | — | — | — | Shared |
| k_damage | — | — | — | +0.520*** | — | +0.133*** | Ageing |
| k_repair_clear | — | — | — | −0.487*** | — | −0.094*** | Ageing |
| k_repair_decay | — | — | — | +0.491*** | −0.689*** | +0.593*** | Ageing |
| k_rapamycin □ | — | — | — | −0.469*** | +0.689*** | −0.631*** | Ageing (KEY) |
| k_BioAgeGap | — | — | — | — | — | −0.416*** | Ageing |

## Data Availability

The SBML model (IQANOVA_GLP1_Ageing_QSP.xml), synthetic calibration dataset (all_synthetic_data.csv), parameter table (IQANOVA_Parameter_Table.xlsx), GSA results (gsa_results.json), Bayesian meta-analysis outputs, and figure-generation scripts are included in the supplementary package accompanying this submission. On acceptance, the SBML/COMBINE archive will be deposited in BioModels and the full reproducibility package will be archived in Zenodo and mirrored at the IQANOVA ATLAS (https://www.iqanova.org). Repository accession numbers and persistent links will be added at proof stage.

## Code Availability

The SBML model passes structural validation with zero errors in libSBML v5.21. Python scripts for calibration, GSA, Bayesian meta-analysis, and identifiability will be released via www.iqanova.org and Gitgub.

## Author Contributions

I.G.: Conceptualization, model development, calibration, formal analysis, writing — original draft, supervision. I.G. (Irina Goryanin): writing — review and editing, supervision. B.D.: writing — review and editing, business context. All authors read and approved the final manuscript.

## Ethics Approval

Not applicable. This study uses published aggregate clinical trial data and mathematical modelling; no human participants or animal subjects were involved.

## References

1. López-Otín C, Blasco MA, Partridge L, Serrano M, Kroemer G. Hallmarks of aging: an expanding universe. Cell. 2023;186:243–278.

2. Ornago AM et al. Shared and specific blood biomarkers for multimorbidity. Nat Med. 2026;32:736–745. 10.1038/s41591-025-04038-2.

3. Li Y et al. Estimated glucose disposal rate and frailty in middle and older aged adults from three prospective cohorts. Sci Rep. 2025.

4. Wilding JPH et al. Once-weekly semaglutide in adults with overweight or obesity (STEP 1). N Engl J Med. 2021;384:989–1002.

5. Davies M et al. Semaglutide 2.4 mg once a week in adults with overweight or obesity and type 2 diabetes (STEP 2). Lancet. 2021;397:971–984.

6. Garvey WT et al. Two-year effects of semaglutide in adults with overweight or obesity (STEP 4/extension). Nat Med. 2022;28:2083–2091.

7. Wanner C et al. Empagliflozin and progression of kidney disease in type 2 diabetes (EMPA-REG OUTCOME). N Engl J Med. 2016;375:323–334.

8. Rena G, Hardie DG, Pearson ER. The mechanisms of action of metformin. Diabetologia. 2017;60:1577–1585.

9. Mannick JB et al. mTOR inhibition improves immune function in the elderly. Sci Transl Med. 2014;6:268ra179.

10. Dalla Man C et al. A model of GLP-1 action on insulin secretion in nondiabetic subjects. Am J Physiol Endocrinol Metab. 2010;298:E1115–E1121.

11. Balazki P, Schaller S, Eissing T, Lehr T. A physiologically based QSP model of incretin hormones GLP-1 and GIP. CPT Pharmacometrics Syst Pharmacol. 2020;9:353–362.

12. Bosch R et al. A novel integrated QSP model of in vivo human glucose regulation. CPT Pharmacometrics Syst Pharmacol. 2022;11:302–317.

13. Clegg A, Young J, Iliffe S, Rikkert MO, Rockwood K. Frailty in elderly people. Lancet. 2013;381:752–762.

14. Fried LP et al. Frailty in older adults: evidence for a phenotype. J Gerontol A Biol Sci Med Sci. 2001;56:M146–M156.

15. Patel S et al. GDF15 as a biomarker of biological ageing and systemic stress. Nat Aging. 2024;4:312–325.

16. Tuttle KR et al. Semaglutide and kidney outcomes in diabetes and CKD (FLOW trial). N Engl J Med. 2023;389:1954–1966.

17. Frías JP et al. Tirzepatide versus semaglutide once weekly in patients with type 2 diabetes. Lancet Diabetes Endocrinol. 2021;9:765–776.

18. Marso SP et al. Semaglutide and cardiovascular outcomes in patients with type 2 diabetes (SUSTAIN 6). N Engl J Med. 2016;375:1834–1844.

19. Lincoff AM et al. Semaglutide and cardiovascular outcomes in obesity without diabetes (SELECT). N Engl J Med. 2023;389:2221–2232.

20. Husain M et al. Oral semaglutide and cardiovascular outcomes in patients with type 2 diabetes (PIONEER 6). N Engl J Med. 2019;381:841–851.

21. Rubino D et al. Effect of continued semaglutide treatment after 20 weeks in patients with obesity (STEP 4). JAMA. 2021;325:1414–1425.

22. Wadden TA et al. Semaglutide as a supplement to intensive behavioral therapy for obesity (STEP 3). JAMA. 2021;325:1403–1413.

23. Wiviott SD et al. Dapagliflozin and cardiovascular outcomes in type 2 diabetes (DECLARE–TIMI 58). N Engl J Med. 2019;380:347–357.

24. McMurray JJV et al. Dapagliflozin in patients with heart failure and reduced ejection fraction (DAPA-HF). N Engl J Med. 2019;381:1995–2008.

25. Perkovic V et al. Canagliflozin and renal outcomes in type 2 diabetes and nephropathy (CREDENCE). N Engl J Med. 2019;380:2295–2306.

26. Neal B et al. Canagliflozin and cardiovascular and renal events in type 2 diabetes (CANVAS). N Engl J Med. 2017;377:644–657.

27. Turner RC et al. Glycemic control with diet, sulfonylurea, metformin, or insulin in patients with type 2 diabetes (UKPDS 49). JAMA. 1999;281:2005–2012.

28. Knowler WC et al. Reduction in the incidence of type 2 diabetes with lifestyle intervention or metformin (DPP). N Engl J Med. 2002;346:393–403.

29. Gelman A. Prior distributions for variance parameters in hierarchical models. Bayesian Anal. 2006;1:515–534.

30. Salvatier J, Wiecki TV, Fonnesbeck C. Probabilistic programming in Python using PyMC3. PeerJ Comput Sci. 2016;2:e55.

31. Higgins JPT et al. Measuring inconsistency in meta-analyses. BMJ. 2003;327:557–560.

32. Saltelli A et al. Global Sensitivity Analysis: The Primer. Wiley; 2008.

33. Brun R et al. Practical identifiability analysis of large environmental simulation models. Water Resour Res. 2001;37:1015–1030.

34. Raue A et al. Structural and practical identifiability analysis of partially observed dynamical models. Bioinformatics. 2009;25:1923–1929.

35. Venzon DJ, Moolgavkar SH. A method for computing profile-likelihood-based confidence intervals. Appl Stat. 1988;37:87–94.

36. FDA (2020). Model-Informed Drug Development: Qualifying New Approaches. FDA Guidance.

37. ICH M15 (2023). General Principles for Model-Informed Drug Development.

38. ASME V&V40 (2018). Assessing Credibility of Computational Modeling through Verification and Validation: Medical Devices.

39. Hucka M et al. The systems biology markup language (SBML). Bioinformatics. 2003;19:524–531.

